# Low TET1 Expression Levels in COPD Are Associated with Airway and Blood Neutrophilia

**DOI:** 10.1101/2025.04.15.25325889

**Authors:** Hong Ji, Xue Zhang, Angela L. Linderholm, Maya Juarez, Michael Schivo, Brooks Kuhn, Richart W. Harper, Amir A. Zeki, Angela Haczku

**Affiliations:** UC Davis Lung Center, University of California, Davis, CA; Department of Anatomy, Physiology and Cell biology; School of Veterinary Medicine, Davis, CA; Division of Human Genetics, Cincinnati Children’s Hospital Medical Center, Cincinnati, OH; Division of Pulmonary, Critical Care, and Sleep Medicine; Department of Internal Medicine, School of Medicine, Davis, CA; Veterans Affairs Medical Center, Mather, CA, USA

## Abstract

Epigenetic dysregulation, particularly DNA methylation variations, is implicated in the pathogenesis of chronic obstructive pulmonary disease (COPD). Ten-eleven translocation (TET) proteins (TET1, TET2, and TET3) regulate DNA methylation and gene transcription. Impaired TET1 expression was previously associated with airway inflammation and asthma. Here we investigated TET gene associations with COPD severity. We found that reduced TET1 expression in peripheral blood mononuclear cells was associated with higher sputum and blood neutrophil counts, decreased lung function and increased disease severity in patients. These findings support a potential protective role and warrant further mechanistic investigations into the actions of TET1 in COPD.

---

Epigenetic dysregulation is an emerging mechanism implicated in chronic obstructive pulmonary disease (COPD). Variations in DNA methylation are found in genes and pathways associated with COPD etiology and severity ^1,2^. While the cause of these variations remains unclear, evidence supports the involvement of Ten-eleven translocation methylcytosine dioxygenase (TET) proteins (TET1, TET2, and TET3) known to catalyze the hydroxylation of 5-methylcytosine and regulate DNA methylation. TET proteins are transcriptional regulators of inflammatory, hematopoietic, and oxidative stress related mechanisms, often with differential function ^3^. Our lab previously showed that TET1 inhibits allergic airway inflammation ^4^, while studies in patients and mouse models of COPD implied TET2 as protective ^5,6^. Interestingly, TET1 expression was reduced in COPD both in the lungs of patients and mouse models ^2,7^ However, the role of TET1 in COPD remains unclear.

Here we examined the associations between the expression of TET family genes with airway neutrophilia, inflammatory and oxidative stress pathways and COPD severity. We used bulk RNA-seq assessment of peripheral blood mononuclear cell (PBMC) samples from a cohort of healthy subjects (N=15) and COPD patients (N=17). Lung function measurements, differential cell counts in blood and induced sputum were also performed on these subjects. Detailed recruitment, sample processing and data acquisition are described in the Supplemental Material and Methods section. COPD patients showed a significantly decreased lung function and elevated neutrophils in the sputum and blood (Supplementary Table 1). Sputum neutrophils (%) negatively associated with lung function in all participants (FEV1, FEV1% predicted, and FEV1/FVC) and in COPD patients (FEV1) (Table 1). Neutrophils positively associated with higher COPD Assessment Test (CAT) scores in all participants (p=0.001) and in COPD patients (p=0.172) (Figure 1A and Table 1). Higher CAT scores indicate greater symptom burden. Blood neutrophil percentage and absolute counts were also negatively associated with lung function and positively associated with CAT in all participants (Table 1).

**Table 1:** Correlations between sputum and blood neutrophil parameters with lung function and CAT.

|  | **Sputum NP%** | **Sputum NP%**<br>**(no squamous cells)** | **PBMC**<br>**neutrophilx10<sup>3</sup>/uL** | **PBMC**<br>**neutrophil absolute** | **PBMC**<br>**Neutrophil%** |
| --- | --- | --- | --- | --- | --- |
| **All participants** |  |  |  |  |  |
| Lung function |  |  |  |  |  |
| FEV1 | **p=0.006**, r= -0.490 | **p=0.0003**, r= -0.664 | **p=0.041**, r= -0.413 | **p=0.020**, r= -0.436 | p=0.053, r= -0.370 |
| FEV1% | p=0.055, r= -0.354 | **p=0.003**, r= -0.575 | **p=0.005**, r= -0.542 | **p=0.003**, r= -0.544 | **p=0.040**, r= -0.391 |
| FEV1/FVC | p=0.051, r= -0.359 | **p=0.0004**, r= -0.652 | **p=0.008**, r= -0.628 | **p=0.0005**, r= -0.612 | **p=0.030**, r= -0.410 |
| CAT | p=0.061, r=0.346 | **p=0.003**, r=0.564 | p=0.098, r=0.331 | **p=0.045**, r=0.376 | **p=0.015**, r=0.449 |
| **COPD patients** |  |  |  |  |  |
| Lung function |  |  |  |  |  |
| FEV1 | p=0.204, r= -0.335 | **p=0.005**, r= **-0.752** | p=0.384 | p=0.361 | p=0.932 |
| FEV1% | p=0.585 | p=0.083, r= -0.520 | p=0.758 | p=0.842 | p=0.728 |
| FEV1/FVC | p=0.910 | p=0.148, r= -0.445 | p=0.511 | p=0.607 | p=0.601 |
| CAT | p=0.587 | p=0.172, r=0.404 | p=0.438 | p=0.442 | p=0.595 |

**Figure 1.**
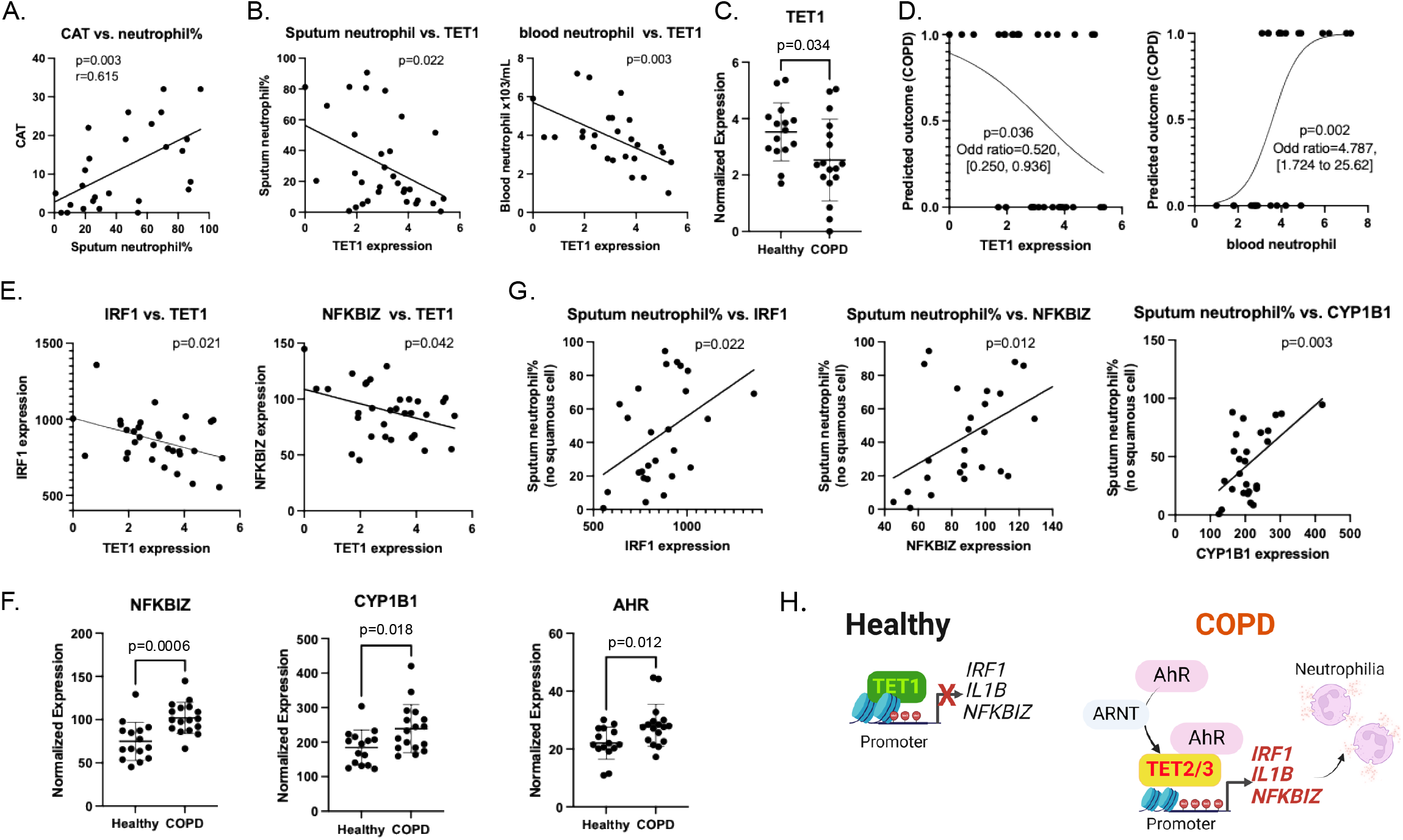
Relationships between Neutrophil counts, blood TET normalized expression values and COPD and implicated mechanisms. **A**. Sputum neutrophil% (after squamous cell removal) positively correlates with CAT scores. **B**. Association between TET1 expression with sputum neutrophils% and blood neutrophils (NEUTROPHILx10^3^/uL). **C**. TET1 expression in PBMC from healthy subjects and COPD patients. **D**. Logistic regression plots showing the prediction of COPD by TET1 expression level and blood neutrophil concentration. Logistic regression models were applied. **E**. Correlation between TET1 and IRF1, NFKBIZ expression levels in PBMC. **F**. Expression levels of NFKBIZ, CYP1B1 and AHR in PBMC from healthy subjects and COPD patients. **G**. Associations of IRF1, NFKBIZ, and CYP1B1 expression levels in PBMC with sputum neutrophil%. **H**. Diagram of proposed model. In healthy subjects, TET1 in PBMCs limits the expression of cytokines and transcription factors, likely by controlling chromatin and histone modification instead of DNA methylation. In COPD patients, TET1 expression is low and TET2 and/or TET3 would replace TET1 and bind to the promoter of these genes, remove DNA methylation and promote their expression. For expression of genes, normalized expression values from RNA-seq are plotted.

We discovered that *TET1* mRNA expression was negatively associated with sputum neutrophil % and blood neutrophil concentration (Figure 1B and Table 2, p=0.080 and p=0.026 after adjusting for age). The negative relationship remained when stratified by COPD status (e.g., p=0.039 for blood NEUTROPHIL x10^3^/uL) in healthy subjects; p=0.178 for sputum neutrophil % in COPD). Consistent with previous observations, there was lower *TET1* expression in the peripheral blood of COPD patients when compared with healthy subjects (Figure 1C). Importantly, age showed no significant contribution (p=0.955) and was excluded from the statistical model. Additionally, *TET1* significantly negatively correlated with *TET3* (p=0.002) while *TET2* and *TET3* expression showed a positive correlation with one another (p=0.026), suggesting differential regulation and function of these genes. Although we did not find statistically significant associations between blood *TET2* or *TET3* expression and COPD, we observed significant positive associations of blood *TET2* and *TET3* with blood neutrophil counts. No significant associations between *TET1, TET2* and *TET3* expression and lung function were found.

**Table 2.**
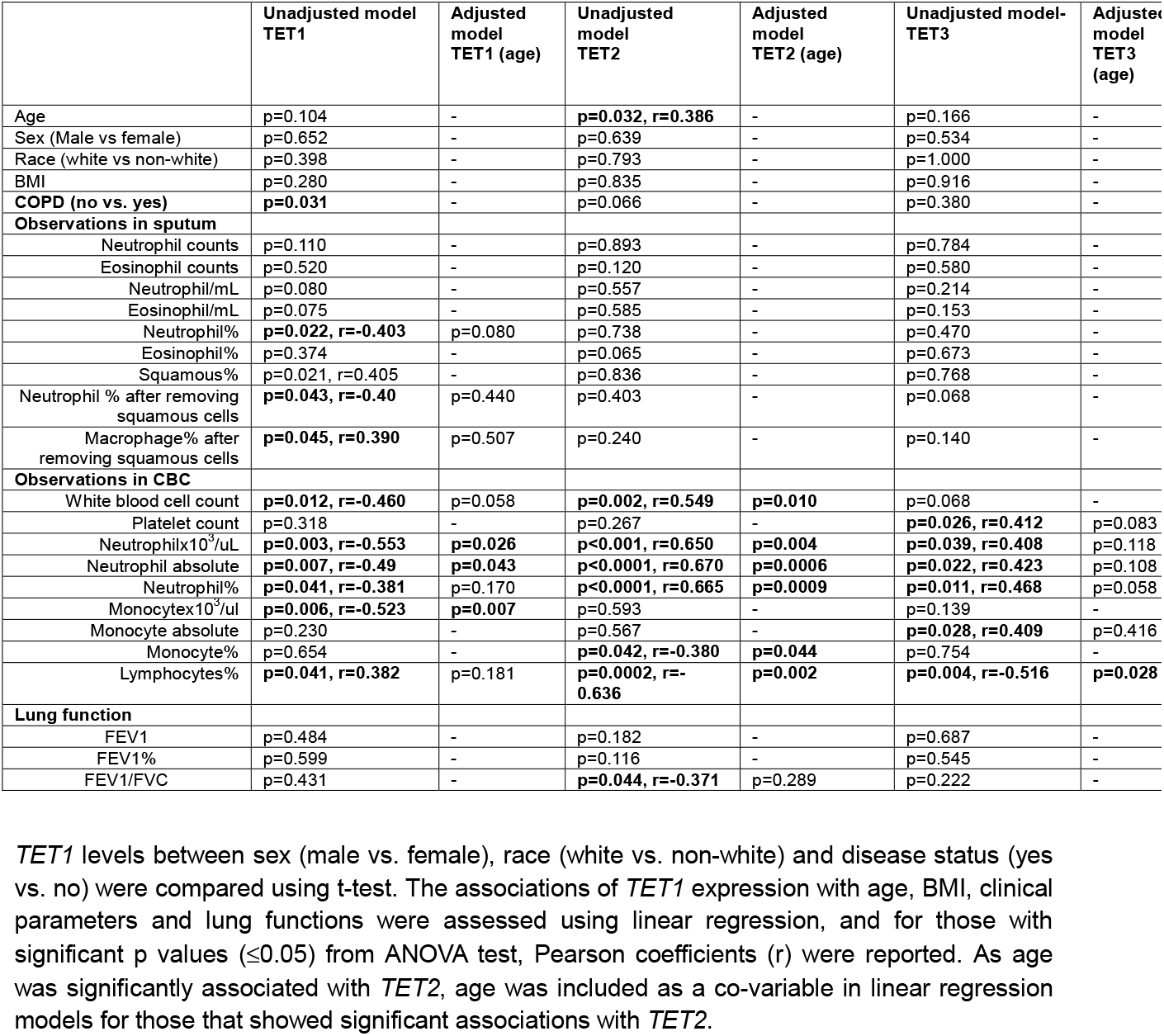
Associations of blood TET mRNA levels with clinical outcomes.

Logistical regression analyses revealed that higher *TET1* expression was associated with a reduced odds of having COPD (odds ratio of 0.520, [95% CI, (0.250, 0.936)] (Figure 1D, p=0.036), suggesting that higher *TET1* expression may be protective. Consistent with the critical role of neutrophils in COPD pathogenesis, we found that higher blood neutrophil counts and sputum neutrophil percentages were associated with an increase in the odds of having COPD [odds ratio=4.78, 95% CI (1.72, 26.62), p=0.002; and odds ratio=1.053, 95% CI (1.018 to 1.102), p=0.004] (Figure 1D). Since sputum neutrophil percent and blood neutrophil counts were also associated with *TET1* expression (Table 2), we used logistic models with and without different neutrophil parameters to investigate whether the impact of *TET1* on COPD may be mediated by neutrophils. We found that the effects (regression coefficients) of *TET1* on COPD decreased when these factors were added to the models as the mediator, suggesting potential mediation by these factors, though none of them reached the statistical significance in the causal inference test (p=0.08-0.11, Supplementary Table 2). These data suggest that TET1 may limit neutrophil counts. While our study sample size is small, as a discovery cohort this information is useful for hypothesis generation and to guide future investigations.

Our prior studies in human airway epithelial cells support a new role of TET1 in limiting cytokine production through histone modification and chromatin accessibility ^8^. The TET proteins may interact with other epigenetic regulators and regulate the expression of genes important for neutrophil function and inflammatory (inflammasome and NF-kB) pathways. TET proteins, additionally, have a complex relationship with the Aryl Hydrocarbon Receptor (AhR), a major regulator of oxidative stress. In fact, AhR activation may induce *TET2* expression through promoter binding and interacting with TET2 as transcriptional co-regulators in immune cell lineage specificity during inflammation ^9^. In support of potential differential functions between TET1 and TET2, we observed significant negative correlations between *TET1* mRNA with *IRF1* and *NFKBIZ* (Figure 1E), but positive correlations between *TET2* with *IRF1* and *NFKBIZ*, and positive correlations between *TET3* with *IRF1* and *NFKBIZ* (Supplementary Figure 1). TET2, *TET3* and *ARNT* also significantly positively correlated with *IL1B* while *TET1* showed a negative trend (p=0.281) (Supplementary Figure 1). Importantly, *IRF1, NFKBIZ, CYP1B1* and *AHR* were upregulated in COPD patients (Figure 1F). Sputum neutrophils (%) were positively associated with *IRF1, NFKBIZ* and *CYP1B1* (a direct target of the AhR receptor) (Figure 1G).

Our study provides intriguing new findings supporting a protective role for TET1 in COPD potentially through limiting neutrophilia. Decreased peripheral blood cell *TET1* expression was associated with higher CAT scores and increased sputum neutrophil percentage. Neutrophils and their products are key contributors to airway inflammation in COPD, and cause many of the disease-associated pathological features, including emphysema and mucus hypersecretion^10^. Reduced *TET1* expression was also associated with an increase of pro-neutrophilic inflammatory genes (e.g., *IL1B, IRF1, NFKBIZ*) (Figure 1H). TET proteins may play differential and possibly opposing roles in regulating the same set of genes in COPD through distinct mechanisms (Figure 1H). Additional mechanistic studies are needed to substantiate these novel observations and decipher how TET1, TET2 and TET3 function in COPD and other chronic airways diseases.

## Declarations

### Ethics approval and consent to participate

This study was reviewed and approved by the University of California, Davis Institutional Review Board.

### Data Availability

All data produced in the present work are available upon reasonable request to the authors.

### Authors Contributions

H.J. and A.H. conceived the study. H.J. performed data analysis with the assistance of X.Z. A.L.L., M.J., M.S., B.K., R.W.H., and A.A.Z. planned and performed clinical studies. H.J., A.A.Z., and A.H. wrote the manuscript.

