## Supplementary Information for "Low TET1 Expression Levels in COPD Are Associated with Airway and Blood Neutrophilia"

**Methods**

**Human subjects**

Patients with COPD (n=17) and non-smoking healthy volunteers (n=15) were recruited from the UC Davis pulmonary and primary care clinics. Adults aged over 18 years with a clinical diagnosis of COPD and a history of smoking (former smokers) were included in the study. Participants with asthma or other lung diseases were excluded. Healthy subjects were non-smokers aged over 18 years, without any known lung disease and other significant lung or major systemic illness that would result in confounding study observations or additional risk to the subject. Subjects that met eligibility criteria were consented and invited to enroll in the study. During a single study visit, the following procedures were conducted: review of medical record, physical examination, pulmonary function testing (PFT), health related quality of life surveys [modified medical research council scale (mMRC) and COPD assessment test (CAT)], venipuncture for collection of 20 mL blood, sputum induction by inhalation of 3% hypertonic saline, and 6-minute walk test. Healthy volunteers underwent all procedures except for the physical examination. For COPD patients, a physical examination, PFT, and 6-minute walk test performed within 2 months of the initial study visit was acceptable. This study was reviewed and approved by the University of California, Davis Institutional Review Board.

**Sputum**

10% Sputolysin reagent (MilliporeSigma, Burlington, MA) was added 1:1 weight/volume to induced sputum specimens. The sample was then placed in a 37°C shaking water bath for 15 minutes. Every 5 minutes, the sample was removed to be pipette mixed. Undiluted Sputolysin reagent was added as necessary if sample homogenization was poor. Sputum cells were strained through a 40 µm Olympus Advanced cell strainer (Genesee Scientific, San Diego, CA) before cell count by the Countess™ Automated Cell Counter. Approximately $32 \times 10^3$ cells were centrifuged at 500 rpm for 5 minutes onto a microscope slide, then stained via the Shandon Kwik-Diff staining kit (Thermo Fisher Scientific, Waltham, MA). 2 cytospins were made for each subject. The remaining cells were apportioned for flow cytometry, while the sputum supernatant was aliquoted and snap frozen for storage at -80°C. Cytospins were evaluated for the presence of macrophages/monocytes (M0), lymphocytes (LC), neutrophils (NP), eosinophils (EP), epithelial cells, and oral squamous cells. At minimum, 200 cells were counted per cytospin and differential cell percentages were averaged between cytospins for each subject. Sputum samples were deemed poor quality if >40% of the cytospin was squamous cells.

**RNA-seq analysis**

RNA was isolated from the cytology brushes using the Qiagen RNeasy Micro Kit (Qiagen, Germantown, MD, USA). cDNA Libraries were prepared with the KAPA Stranded RNA-Seq Kit with the RiboErase Kit (Roche, Pleasanton, CA, USA) according to the manufacturer's protocol. Generation of cDNA libraries and sequencing was carried out by the DNA Technologies and Expression Analysis Core Laboratories at the University of California Davis Genome Center. Genes with <0.5 counts per million reads in all samples were filtered prior to the analysis. Data were transformed using a variance-stabilizing transformation and then LOESS (locally estimated scatterplot smoothing) normalized. Gene annotation was based on Gencode genome assembly version GRCh38.

**Statistical Analysis**

Prior to statistical analysis, data quality and distributions were examined. To compare the demographics and characteristics between subjects with and without COPD, we conducted two sample t tests and Fisher’s exact tests according to the data distribution, using SAS 9.4. The correlation among expression of TET1, 2 and 3 was assessed with Pearson’s correlation using JMP Pro 17. The association between TET expression and other parameters were assessed either using linear regression (for continuous data), ANOVA or student t test (categorical data). To test the association of COPD with cytokines, we performed logistic regression (SAS 9.4 and JMP Pro 17). To test if the effect of TET1 on COPD may be mediated by the cytokines, we examined the change in TET1 effect when individual factors was included or excluded from the modeling. Mediation was also tested using R package ‘cit’ [1,2]. $p<0.05$ was considered statistically significant.

**Supplementary Figure 1. Correlations between expression of TET, HDAC2 and genes involved in neutrophil recruitment and function.**

Pearson correlation between normalized expression levels from RNA-seq between **A**) *TET2* and *IRF1*, *TET2* and *NFKBIZ*, *TET2* and *IL1B*, **B**) *TET3* and *IRF1*, *TET3* and *NFKBIZ*, *TET3* and *IL1B*, and **C)** *ANRT* and *IL1B*, *TET1* and *IL1B*, and *HDAC2* and *IRF1*.

**Supplementary Table 1. Demographics of subjects included in this study.**

| Variable | COPD (N=17) | healthy (N=15) | *P* Value |
|---|---|---|---|
| Age, median (Q1, Q3) | 70.0 (64.0, 77.0) | 35.5 (27.0, 46.0) | <0.001 |
| Sex: (M), n (%) | 11 (65%) | 6 (43%) | 0.29 |
| Race, n (%) | | | 0.85 |
| . Asian | 2 (13%) | 0 (0%) | |
| . Black | 2 (13%) | 1 (11%) | |
| . White | 10 (67%) | 8 (89%) | |
| . White/ American Indian | 1 (7%) | 0 (0%) | |
| BMI, median (Q1, Q3) | 25.0 (21.0, 37.0) | 23.0 (21.0, 24.0) | 0.29 |
| **Observations in sputum** | | | |
| Squamous%, median (Q1, Q3) | 26.8(4.1, 49.5) | 29.7(27.5, 46.6) | 0.23 |
| Neutrophil%, median (Q1, Q3) | 29.1(19.4, 69.1) | 13.1(5.5, 18.8) | 0.013 |

| | | | |
|---|---|---|---|
| Neutrophil% after removing squamous cells, median (Q1, Q3) | 69.1(46.1, 82.8) | 22.0 (10.4, 29.1) | 0.004 |
| Neutrophil count, median (Q1, Q3) | 135.0 (56.0, 313.0) | 42.0 (14.0, 135.0) | 0.009 |
| Neutrophil/mL, median (Q1, Q3) | 559687 (315972, 1610000) | 195665 (83881, 523052) | 0.022 |
| Eosinophil%, median (Q1, Q3) | 0.7 (0.2, 2.2) | 0.0 (0.0, 0.0) | <0.001 |
| Eosinophil% after removing squamous cells, median (Q1, Q3) | 1.0(0.3, 2.1) | 0.0(0.0, 0.0) | <0.001 |
| Eosinophil count, median (Q1, Q3) | 4.0 (1.0, 11.0) | 0.0 (0.0, 0.0) | <0.001 |
| Eosinophil/mL, median (Q1, Q3) | 7214.4 (2523.8, 62066) | 0.0 (0.0, 0.0) | 0.001 |
| Macrophage % | 19.2± 15.8 | 37.0± 19.2 | 0.007 |
| Macrophage % after removing squamous cells | 25.4± 18.9 | 62.3± 22.1 | <0.001 |
| Macrophage count, median (Q1, Q3) | 83.0(33.0, 111.0) | 118.0(53.0, 208.0) | 0.09 |
| Macrophage/mL, median (Q1, Q3) | 265333(138809, 609375) | 545747(340099, 912734) | 0.05 |
| Lymphocytes%, median (Q1, Q3) | 1.4(0.7, 2.0) | 0.7(0.0, 1.5) | 0.027 |
| Lymphocytes% after removing squamous cells, median (Q1, Q3) | 1.6(1.0, 2.6) | 1.0(0.3, 2.1) | 0.19 |
| Lymphocytes count, median (Q1, Q3) | 6.0(4.0, 9.0) | 3.0(0.0, 5.0) | 0.008 |
| Lymphocytes/mL, median (Q1, Q3) | 29085(15143, 43286) | 11036(3507.7, 21587) | 0.034 |
| SP-D, median (Q1, Q3) | 21861(11300, 32428) | 1630.0(1191.3, 4490.7) | 0.001 |
| **Observations in CBC** | | | |
| White blood cell count | 7.4±1.7 | 5.2±0.9 | <0.001 |
| Platelet count | 243.0± 55.2 | 216.3± 39.7 | 0.16 |
| Neutrophil%, median (Q1, Q3) | 64.7(60.8, 67.1) | 58.6(44.6, 62.0) | 0.012 |
| Neutrophilx103/uL | 4.7±1.4 | 2.9±1.1 | 0.001 |
| Neutrophil Absolute | 4.7±1.2 | 2.9±1.0 | <0.001 |
| Monocyte% | 8.6±2.5 | 8.4±2.4 | 0.86 |
| Monocytex103/uL | 0.6±0.1 | 0.4±0.1 | 0.017 |
| Monocyte Absolute, median (Q1, Q3) | 0.6(0.5, 0.7) | 0.4(0.3, 0.5) | 0.004 |

| | | | |
|---|---|---|---|
| Lymphocyte%, median (Q1, Q3) | 23.1(20.7, 27.3) | 29.3(28.5, 43.1) | 0.002 |
| Lymphocytex103/uL | 1.7±0.7 | 1.7±0.4 | 0.96 |
| Lymphocyte Absolute | 1.8±0.6 | 1.8±0.4 | 0.93 |
| SP-D ng/mL, median (Q1, Q3) | 23.3(16.9, 58.9) | 3.1(1.3, 6.8) | <0.001 |
| **Lung function** | | | |
| FEV1/FVC, median (Q1, Q3) | 55.9(48.5, 61.5) | 80.0(78.3, 83.3) | <0.001 |
| FEV1/FVC following bronchodilator, median (Q1, Q3) | 56.0(50.7, 65.1) | 82.5(80.2, 87.2) | <0.001 |
| % Change FVC, median (Q1, Q3) | 4.5(0.8, 7.5) | 0.6(-0.3, 1.9) | 0.049 |

Sex and race were tested using Fisher's exact test. Variables shown as mean±SD were tested using ANOVA. Variables shown as median(IQR) were tested using Kruskal-Wallis test.

**Supplementary Table 2. Models and Causal inference test (CIT) to evaluate whether neutrophil parameters and SP-D mediate the association of disease with TET1.**

| | **Model** | **β** | **P** | | | |
|---|---|---|---|---|---|---|
| | Disease=TET1 | -0.6534 | 0.047 | | | |
| | | | | | | **CIT** |
| Mediator (M) | model | β (TET1) | P (TET1) | β (M) | P (M) | P (cit) |
| sputum Neutrophil% | M=TET1 | -8.5574 | 0.022 | | | **0.111** |
| | Disease= TET1+M | -0.4883 | 0.166 | 0.0304 | 0.09 | |
| Blood Neutrophilx103/uL | M=TET1 | -0.5931 | 0.003 | | | **0.080** |
| | Disease= TET1+M | -0.5068 | 0.295 | 1.2508 | 0.063 | |
| Blood Neutrophil Absolute | M=TET1 | -0.5184 | 0.007 | | | **0.101** |
| | Disease= TET1+M | -0.3363 | 0.428 | 1.7197 | 0.018 | |
| Blood Neutrophil% | M=TET1 | -2.9262 | 0.041 | | | 0.257 |
| | Disease= TET1+M | -0.5844 | 0.129 | 0.1190 | 0.073 | |
| sputum SP_D | M=TET1 | -7624.69 | 0.033 | | | 0.189 |
| | Disease= TET1+M | -0.3573 | 0.369 | 0.000091 | 0.047 | |

For all factors shown here, they all significantly associated with TET1 [in the model M=TET1, p(TET1)<0.05]. In addition, in the model disease=TET1+M, beta (TET1) became smaller when compared to the model with TET alone (in the $2^{nd}$ row, β=-0.6534) and p (TET1) became insignificant. These data suggest mediation even though the formal CIT test on mediation may give a >0.05 p value.
