## Supplementary figures and images for "Low TET1 Expression Levels in COPD Are Associated with Airway and Blood Neutrophilia"

### Supplementary Figure 1

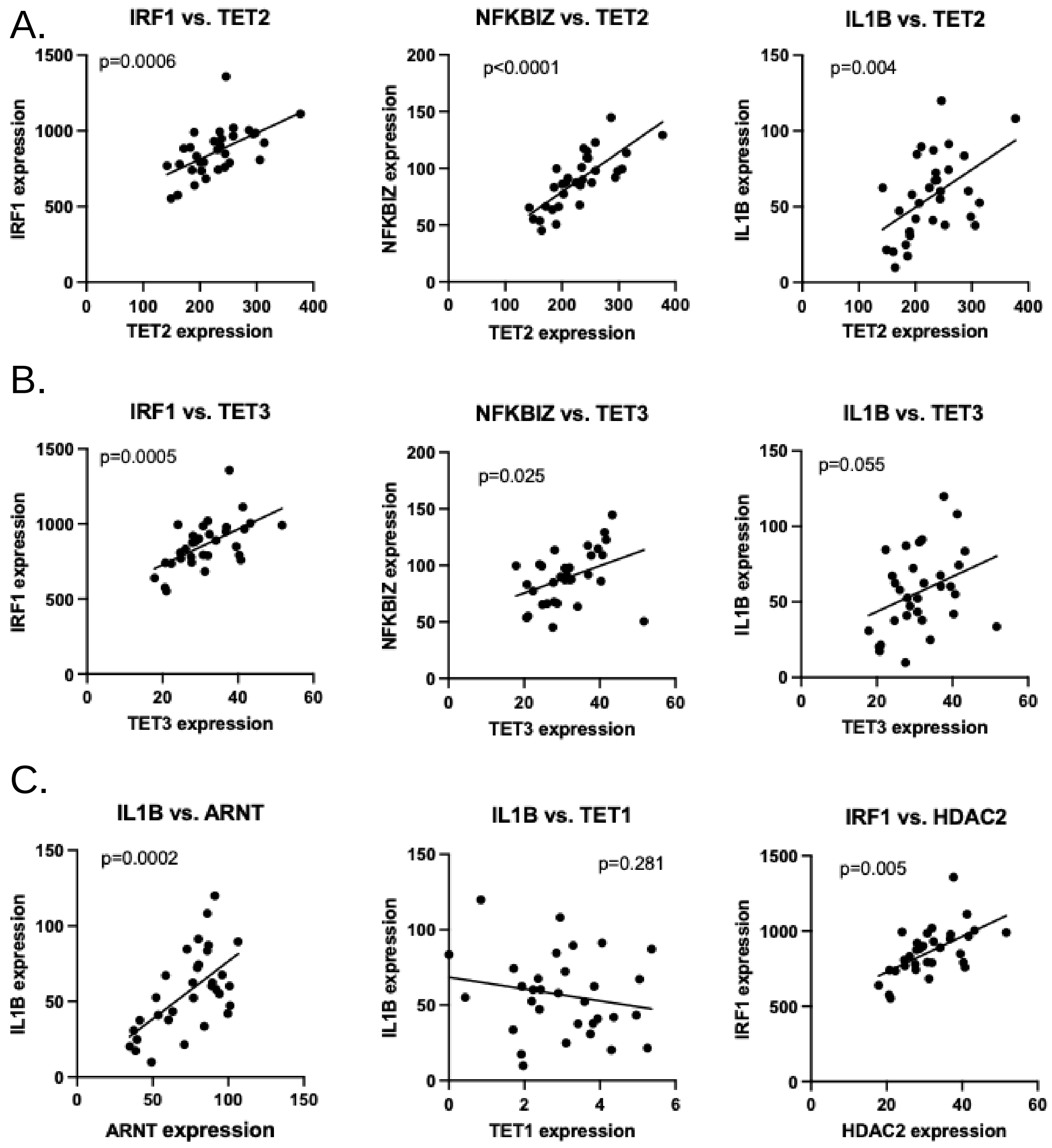


**Supplementary Figure 1**
